# Quantifying the Uncertainty of Human Activity Recognition Using a Bayesian Machine Learning Method: A Prediction Study

**DOI:** 10.1101/2023.08.16.23294126

**Authors:** Hiroshi Mamiya, Daniel Fuller

**Affiliations:** Department of Community Health and Epidemiology, University of Saskatchewan, Rm 3247 - E wing - Health Sciences, 104 Clinic Place, Saskatoon, Saskatchewan, Canada S7N-2Z4

## Abstract

**Background:** Machine learning methods accurately predict physical activity outcomes using accelerometer data generated by wearable devices, thus allowing the investigation of the impact of built environment on population physical activity. While traditional machine learning methods do not provide prediction uncertainty, a new method, Bayesian Additive Regression Trees (BART) can quantify such uncertainty as posterior predictive distribution. We evaluated the performance of BART in predicting physical activity status.

**Methods:** We applied multinomial BART and the benchmark method, random forest, to accelerometer data in 25,424 time points, which were generated by wearable devices attached to 37 participants. We evaluated prediction accuracies and confusion matrix using leave-one-out cross-validation.

**Results:** BART and random forest demonstrated comparable accuracies in prediction.

**Conclusions:** BART is a relatively novel ML method and will advance the incorporation of predicted physical activity status into built environment research. Future research includes the evaluation of the association between the built environment and predicted physical activity with and without accounting for prediction uncertainty.

## Introduction

The application of Machine Learning (ML) is expected to increase in Physical Activity (PA) epidemiology (1,2). ML enables the automated measurement of PA intensity or type (e.g., sitting, walking, or vigorous physical activity), where predictors are accelerometer signals generated by wearable devices such as smartphones. ML can capture complex non-linear functions of a large number of accelerometer variables and has consistently shown superior accuracy in predicting PA categories to traditionally used rule-based classification methods for PA categories (1, 3).

The improved accuracy of predicting PA categories by ML will advance built environment and healthy cities research, which examines the etiologic association between the predicted PA outcome and daily exposure to the modifiable environmental drivers of PA, including walkability and green space (4). One of the overlooked limitations of utilizing ML-predicted variables in public health research is the lack of prediction uncertainty i.e., the lack of confidence interval around the predicted probability of PA status (5–7). Commonly used ML methods to predict PA types, such as random forest, do not provide uncertainty estimates but simply report the most likely PA type as if there is no measurement error. Treating ML-predicted variables without uncertainty leads to underestimated standard errors of the estimated etiologic association between built environmental characteristics and ML-predicted PA categories, which can lead to a falsely conclusive association i.e., higher type I error (6,8).

Bayesian Additive Regression Trees (BART) is a relatively new ML method with growing applications in epidemiology (9,10). BART provides prediction uncertainty in the form of the probability distributions of each of PA activity types to be classified (11,12). Such predictive distribution can then be used to generate multiple predicted values through Monte Carlo sampling, which are then analyzed as if multiply imputed values within the second-stage analysis that estimates the impact of built environment on the predicted PA outcomes (6,13). Thus, BART will effectively allow propagating prediction uncertainty of PA into built environment research.

BART has not been used in PA research to date. Thus, the objective of this study is to compare the accuracy of predicting categories representing PA intensity and types using BART and random forest. The latter is a commonly used ML method to predict PA using accelerometer data (1).

## Methods and data

Our data consisted of accelerometer data containing n=25,424 time points with 5 seconds interval from 37 research participants (mean number of time points: 687, range: 664-723 points per participant). The participants performed the predefined sequences and length of the six combinations of PA types and intensities on a treadmill. These outcome categories are: lying down, sitting, self-paced walking, walking at 3 METs (Metabolic Equivalents of Task), 5 METs, and 7 METs, the latter three intensity categories aligning with the definition of light, moderate, and vigorous PA (14) (Supplementary Figure 1). This study was approved by the Memorial University Interdisciplinary Committee on Ethics in Human Research (ICEHR 20180188-EX). Accelerometer data were collected by Ethica Data app installed in Samsung Galaxy S7 (SM-G930W8) smartphone (15) placed in the right pants or shorts pocket of the participants. Ground truth data for activity types and intensity (i.e., METs) was measured using an Oxycon Pro metabolic cart (Oxycon Pro, Jaeger, Hochberg, Germany) (16). From accelerometer data, we computed 58 predictive features in accordance with the previous studies (17,18) at each 5-seconds time period.

BART is a nonparametric sum-of-tree (additive) ensemble machine learning method, where the contribution of each decision tree to prediction is kept weak by regularization priors to prevent overfitting (12). The procedure to fit BART is provided in Appendix 1. We fitted multinomial BART using the BART package in R software (19). Fitting of the benchmark ML method, random forest, followed our previous papers using the caret package in R software (17,18). Codes and data are publicly available (20,21).

The performance evaluation of BART and random forest followed leave-one-out-cross-validation, where algorithms were fitted to the data from 36 participants, followed by prediction where the fitted algorithms were applied to test data from the remaining one person. After repeating the process 37 times for all participants, we combined the predicted outcomes from the 37 test datasets to compute the metrics for model performance in relation to the ground truth.

The evaluation metric is accuracy specific to each PA category, which is the sum of time points with true positive and true negative status (numerator) divided by all time points (denominator). We also created a confusion matrix and calibration plots. Unlike previous activity prediction research, we also computed accuracy across participants’ self-reported gender identity (female and male) for BART, given the potential heterogeneity of prediction accuracy and uncertainty across gender (22).

## Results

Accuracy of the prediction by BART and random forest were similar (Table 1), and both showed the lowest accuracy for identifying self-paced walking. Confusion matrix (Figure 1) indicates that BART tended to misclassify laying from sitting and self-paced walking from walking at 3METs. Calibration plots also suggest lower goodness-of-fit for self-paced walking and walking at 3METs (Supplementary Figures 2 B and C). While accuracy and confusion matrix provide overall prediction error of BART and random forest in the entire test data, BART also informs about pointwise prediction uncertainty for each sample unit (time point). For example, results from one time point (Supplementary Figure 3) illustrate a largely overlapping posterior predictive probability distribution of self-paced walking and walking at 3 METs, indicating the challenge in classifying two PA categories. This is because participants chose an average self-paced walk intensity of 2.7 METS, making it challenging for the algorithms to distinguish between these intensities. This is also indicated by the corresponding MCMC traces at the same time point (Supplementary Figure 4). Accuracy across participants was highly variable, ranging from 47% to 94% (Supplementary Figure 5). There were noticeable variations in accuracy across genders, but both genders showed the lowest accuracy for self-paced walking (Table 2).

**Table 1.** Comparison of accuracy between random forest and Bayesian Additive Regression Model.

| Class | Random Forest | Bayesian Additive Regression Tree |  |
| --- | --- | --- | --- |
|  |  | Mean | 95% CI |
| Lying | 0.8 | 0.72 | (0.68, 0.75) |
| Sitting | 0.59 | 0.64 | (0.60, 0.69) |
| Walking | 0.37 | 0.48 | (0.42, 0.54) |
| Running (3METs) | 0.66 | 0.6 | (0.55, 0.66) |
| Running (5METs) | 0.75 | 0.69 | (0.64, 0.73) |
| Running (7METs) | 0.78 | 0.74 | (0.71, 0.78) |

95% CI: 95% Credible Interval

**Table 2.**
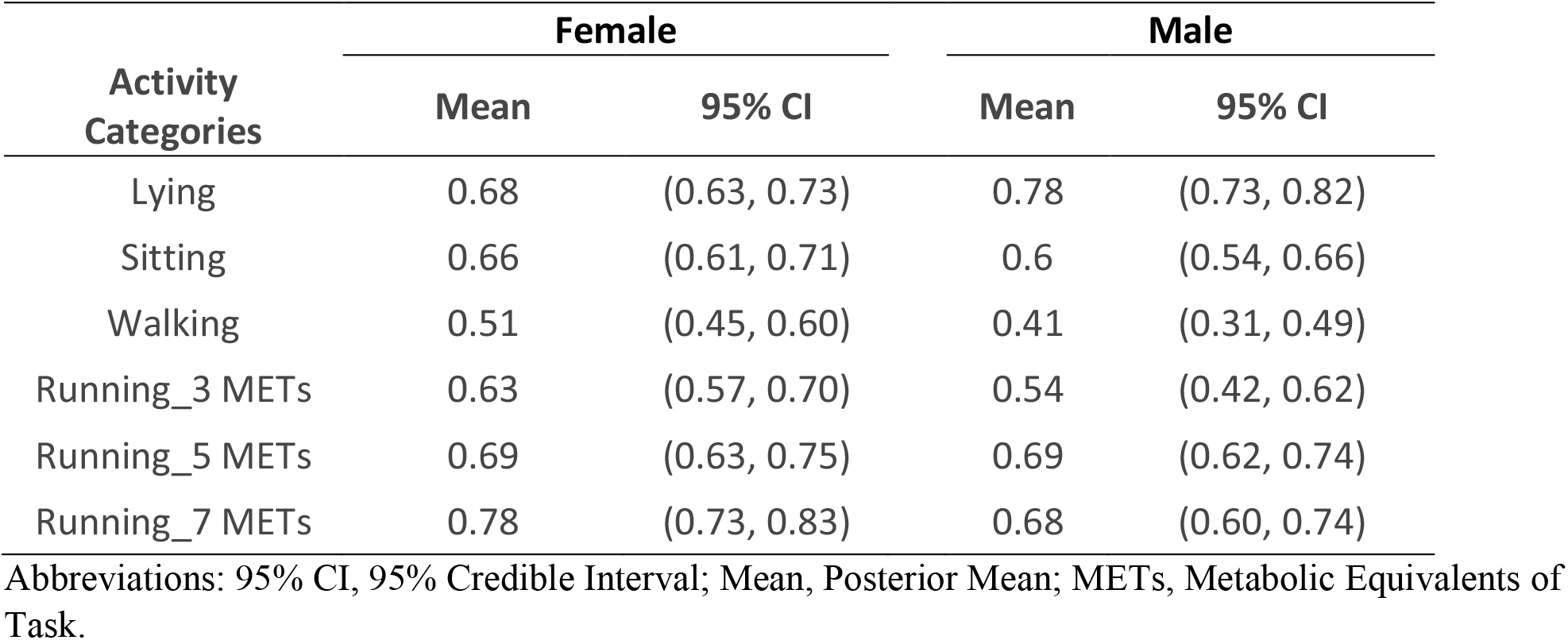
Comparison of accuracy between Male and Female participants by Bayesian Additive Regression Tree.

**Figure 1.**
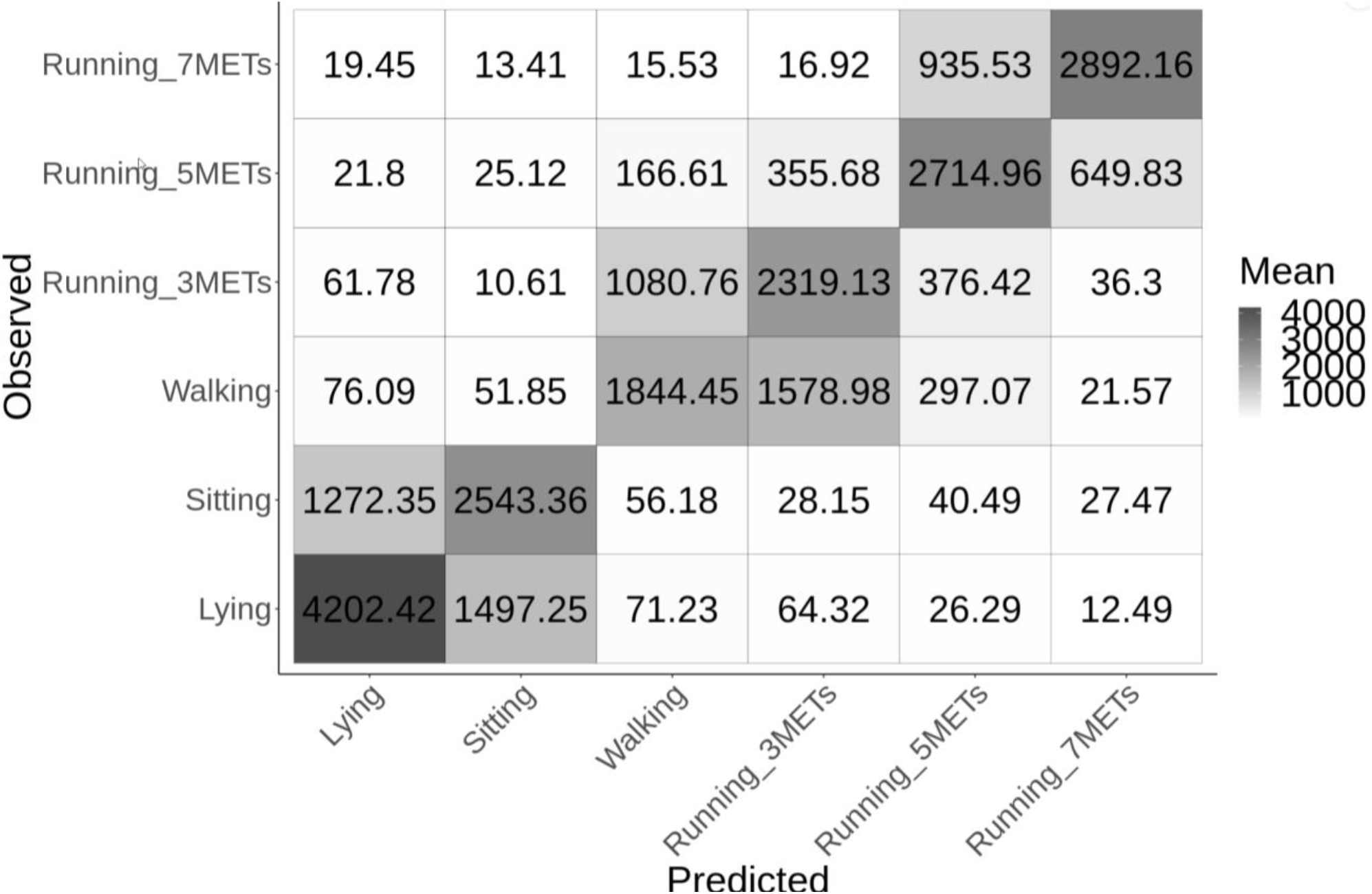
Confusion matrix indicating the posterior mean of the frequency of agreement between the reference (directly observed) and classified (predicted) activity types by Bayesian Additive Regression Tree. Numbers in non-diagonal elements indicate the estimated frequency of misclassification. Abbreviation: METs, Metabolic Equivalents of Task.

## Discussions

The measurement of PA is likely to shift from traditional rule-based methods to ML-based methods due to the accuracy of the latter approach, thereby increasing the validity of built environmental research to examine the impact of built environment on population physical activity (2,23). However, the inability of traditionally used ML algorithms to quantify prediction uncertainty have received scarce attention to date (23,24), and the use of predicted variables in another analysis (e.g., built environment research) without incorporating prediction uncertainty can lead to falsely conclusive association (6). We demonstrated a novel approach to capture such uncertainty using BART, a Bayesian ML that, to our knowledge, has not been utilized in PA research.

BART showed comparable classification accuracy to random forest, a widely used ML for the prediction of PA. However, both algorithms showed lower accuracies than reported in previous studies (1). The lower accuracies maybe due to the wide age variation of participants in our data that may have led to a larger variation of accelerometer noises (25,26). As well, many extant studies used split-sample or k-fold cross-validation that partition data from the same participants into training and test data, rather than using leave-one-out-cross-validation at the participant level, thus reporting inflated accuracy estimates due to leakage issue(27,28). Finally, our study used both PA intensity (various levels of METs) and type as categories for the outcome variable, while the majority of studies used only PA types (1). These make direct comparison of our results to previous research challenging and signifies the importance of standardized evaluation of ML algorithms on publicly accessible accelerometer data in the future (29). While our example is multi-category (multinomial) prediction, BART is readily applicable to predict binary and continuous outcome, for example continuous PA intensity measured by METs.

BART allows measuring the uncertainty of the prediction of PA, as seen in the width and overlap of posterior predictive distributions across categories. This is unlike traditionally used ML algorithms that do not provide interval estimates or predictive distribution but only report one most likely PA type, as if the predicted status is true without potential prediction error. These probability distributions captured by BART readily propagate prediction uncertainty into the second-stage epidemiologic analysis incorporating the ML-predicted PA variables, in a manner similar to probabilistic bias correction or multiple imputation approach (30). BART also potentially overcomes solutions to capture uncertainty in conventional ML using bootstrapping (7,24), which is likely to be computationally expensive, as accelerometer data rapidly becomes large.

As a limitation, the current implementations of BART show challenge in the convergence of the parameter estimation technique, Markov Chain Monte Carlo (MCMC), as the number of observations (hundreds of thousands) and predictors increases (9). Hence, we had to minimize the sample size by aggregating accelerometer signals into 5 second intervals. Since BART demonstrated a comparable accuracy with random forest, we plan to further assess its predictive performance in other publicly available datasets including accelerometer data generated from free-living (non-laboratory) conditions (29), followed by the epidemiologic investigation of association between the exposure to the built urban environment and the predicted PA variable, with and without accounting for uncertainty. Such work will not only promote the quality of evidence in the impact of urban environment, but also advance epidemiologic methodology in incorporating ML-predicted variables. Finally, we caution against the use of socio-demographic and economic status as the predictor, since these personal characteristics are often used as a confounder or effect measure modifier in the second-stage etiologic analysis.

## Data Availability

The de-identified accelerometer data are available publicly at: https://doi.org/10.7910/DVN/LXVZRC.

## Declarations

### Ethics approval and consent to participate

This study was approved by the Memorial University Interdisciplinary Committee on Ethics in Human Research (ICEHR 20180188-EX).

### Consent for publication

Consent for publication was obtained from the study participants.

### Availability of data and materials

Codes to prepare data and fit and evaluate machine learning algorithms are available publicly at the following address: https://github.com/hiroshimamiya/BART_PhysicalActivity. The de-identified accelerometer data are available publicly at: https://doi.org/10.7910/DVN/LXVZRC.

### Competing interests

None

### Funding

The research was supported by the Postdoctoral fellowship from the Artificial Intelligence for Public Health (AI4PH) training platform, Canadian Institute for Health Research. The founder had no role in the design of the study and collection, analysis, and interpretation of data and in writing the manuscript.

### Authors’ contributions

Hiroshi Mamiya conceived the study, designed analytical protocol, and written the manuscript. Daniel Fuller provided inputs in analytical design, collected data, and critically reviewed the manuscript.

## Acknowledgements

We thank SeyedJavad Khataeipour for providing codes to fit and evaluate the random forest algorithm.

## Abbreviations

BART: Bayesian Additive Regression Trees
CI: Credible Interval
ML: Machine Learning
PA: Physical Activity

## Supplementary Digital Contents

**Appendix 1:** Description of Bayesian Additive Regression Tree

Bayesian Additive Regression Trees (BART) is a nonparametric ensemble machine learning algorithm utilizing a sum of regression trees. Rather than relying on a single complex prediction model, BART combines multiple regression trees that provide good predictive performance as a whole. The trees are kept simple (allowed to have only a few branches) to prevent over-fitting, so that optimal out-of-sample prediction performance is achieved. To model our multinomial outcome containing six physical activity categories, we used the BART package in R software (1). Specifically, the package uses probit model to estimate the conditional probability of a response category *j* as:

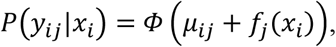

for *j* = {1. . . *K* − 1}, *K* = 6 outcome categories at *i*-th time point in accelerometer data. The function Φ represents the standard normal cumulative density function, with μ_*ij*_ representing intercept, and *f*_*j*_(*x*_*i*_) representing the sum-of-tree function for category *j*, whose predictive features calculated from accelerometer signals at *i-*th time points are denoted *x*_*i*_.

The general form of the ensemble BART function is represented as the sum of *m* trees as

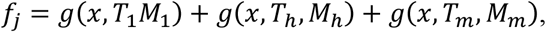

where *T*_*h*_represents tree sizes and *M*_*h*_represents the collection of leaf parameters as:

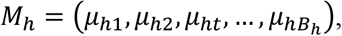

for the total of *B*_*h*_leaves on the *h*th regression tree.

As BART is a fully-Bayesian model, prior probabilities for these parameters need to be specified. Prior on tree depth imposes weak learning (trees with shallow depth), which is accomplished by assigning the probability of a node *d*being non-terminal as:

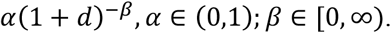

We used the recommended values of α = 0.95 and β = 2, such that the probability of a given tree having a complicated structure (large number of branches) is kept low (2).

The centered prior for the *t-*th leaf parameter covers a large probability range, (Φ[−3.0], Φ[3.0]), and is specified as:

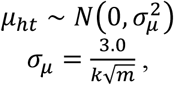

with larger *k* and *m* inducing a stronger shrinkage of the leaf parameters towards zero. We used *k=2* and *m=50* tees for our model as suggested (1).

To estimate the posterior distribution of parameter vector *y*_*ij*_, we ran 2,000 burn-in iterations, followed by 2,000 samples for inference with thinning at every 100 iterations. From the three time-series of accelerometer signals that represent X, Y, and Z axis, we generated 58 predictive features described previously (3).

Accuracy metric of multi-category (as opposed to binary) prediction represents the proportion of agreement between the predicted and observed (i.e., ground truth) categories. We computed this category-specific accuracy metric for each of the 2,000 MCMC interactions, which were averaged over as the posterior mean of class-specific accuracy. Note that the predicted category for each time point was determined as the category with the highest estimated predicted probability, that is, 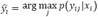 For instance, the predicted category at *i*-th time point is ŷ_*i*_ = *2* when category-specific probabilities at this time point are (0.1, 0.5, 0.1, 0.1, 0.1, 0.1). It is possible to use these probability outputs from BART to calculate accuracy e.g., Brier score. However, we converted these probabilities into the categorical measure for comparison purpose, as the latter measure is the standard output from random forest.

**Supplementary Figure 1.**
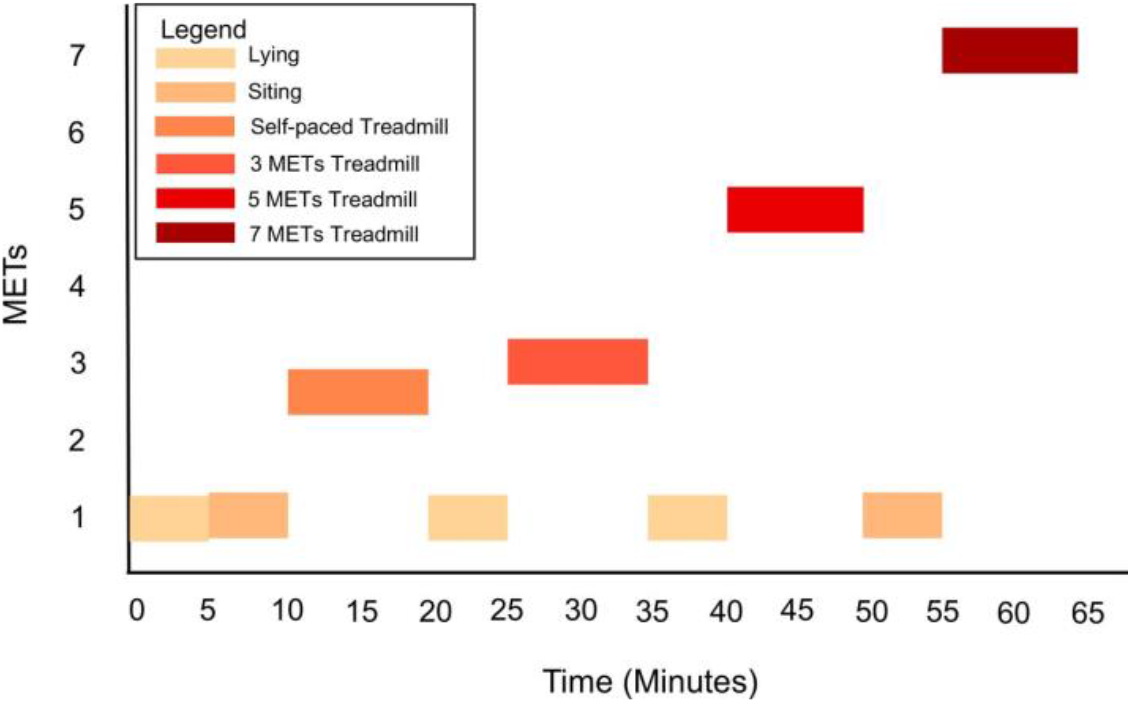
Standardized activity protocol, illustrating the temporal transition of activity types and intensities performed by participants. Abbreviation: METs, metabolic equivalents of task.

**Supplementary Figure 2.**
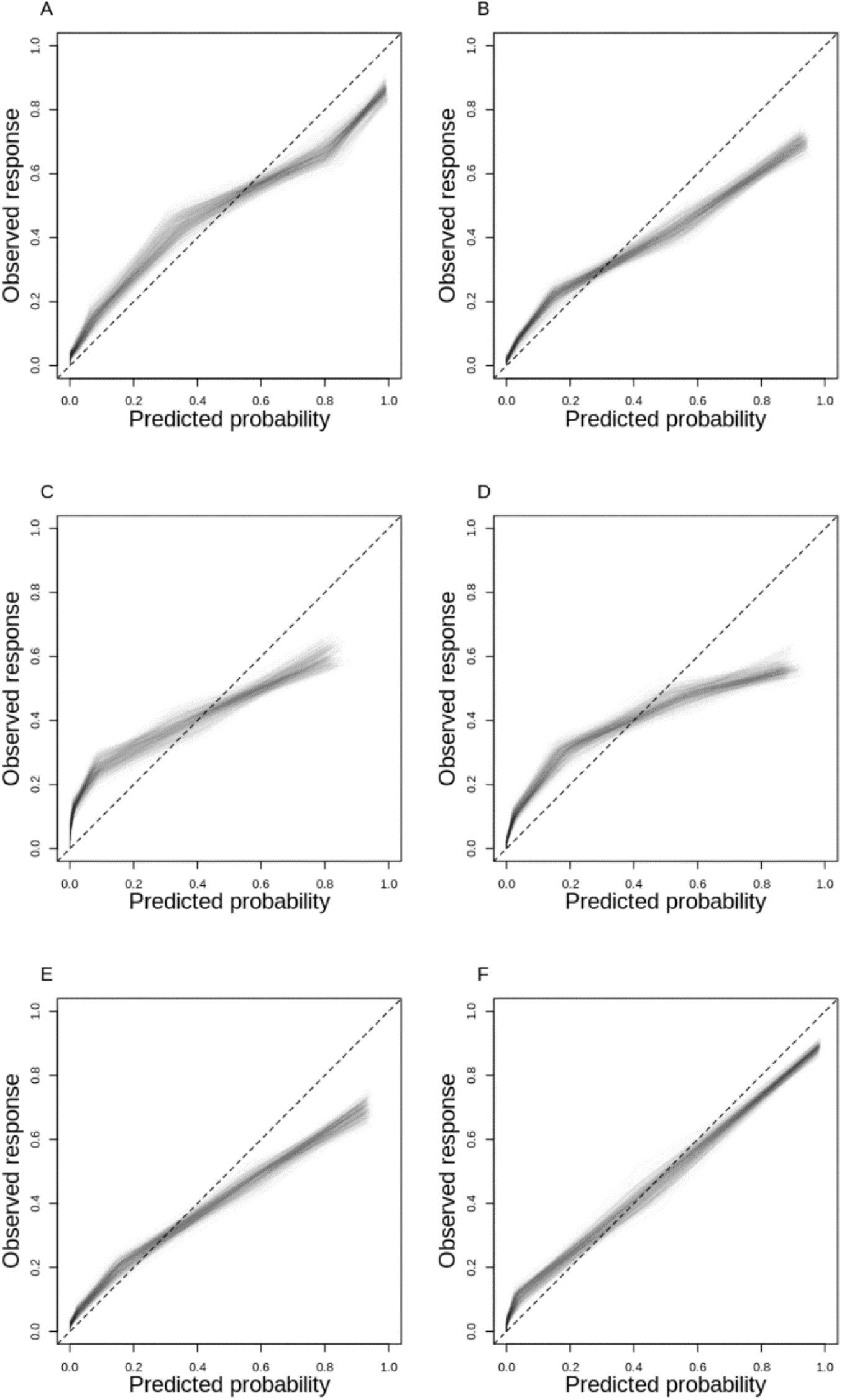
**A-F**: Calibration plots of Bayesian Additive Regression Trees (BART) showing the goodness-of-fit between the decile of predicted probabilities and the proportion of outcomes that are: A) lying, B) sitting, C) self-paced walking, D) running at 3 METs, E) running at 5 METs, and F) running at 7 METs. Translucent lines represent the iterations of Markov Chain Monte Carlo. The x-axis indicates the decile of predicted probability, and y axis indicates the proportion of the outcome corresponding to the deciles of predicted probabilities. Panel C and D shows a larger miscalibration (discordance) between the predicted and observed outcome probabilities, with BART underestimating the outcomes when predicting low probabilities, and overestimating when predicting high probabilities. On the other hand, BART generated predictive probabilities that closely match to the observed frequency when predicting Running 7 METs (Panel F).

**Supplementary Figure 3.**
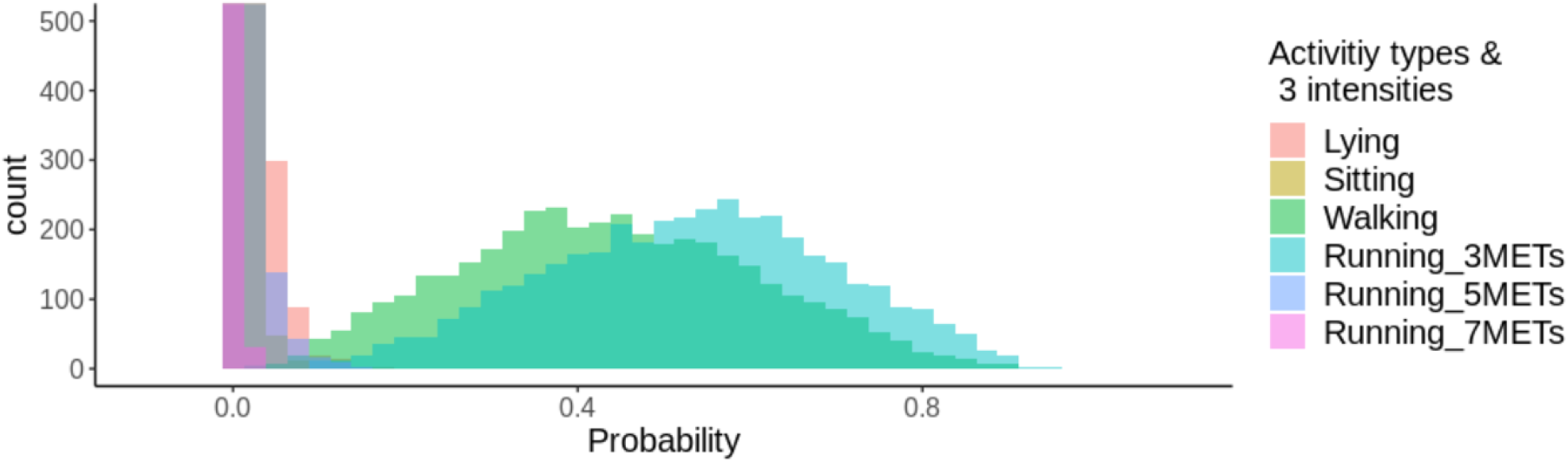
Posterior predictive distribution of six activity categories at one time point, where the estimated distributions of walking and Running (3METs) are largely overlapping. Abbreviation: METs, Metabolic Equivalents of Task.

**Supplementary Figure 4.**
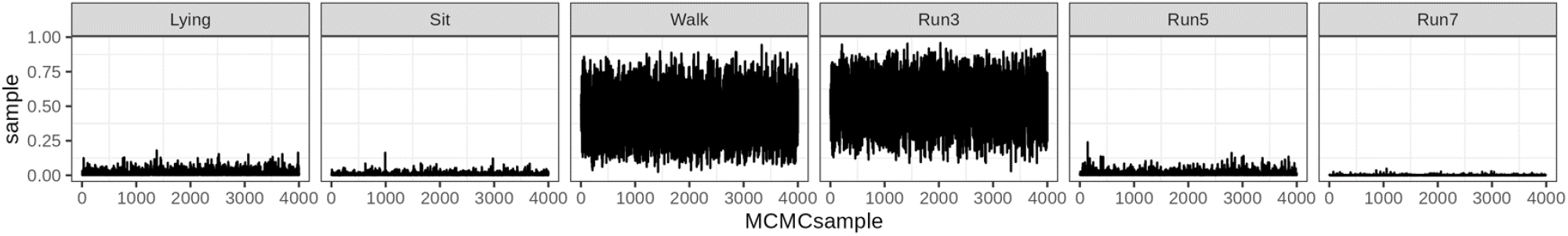
Trace plots of a time point corresponding to Supplementary Figure 3 above, indicating the mixing of Markov Chain for each of the six activity categories.

**Supplementary Figure 5.**
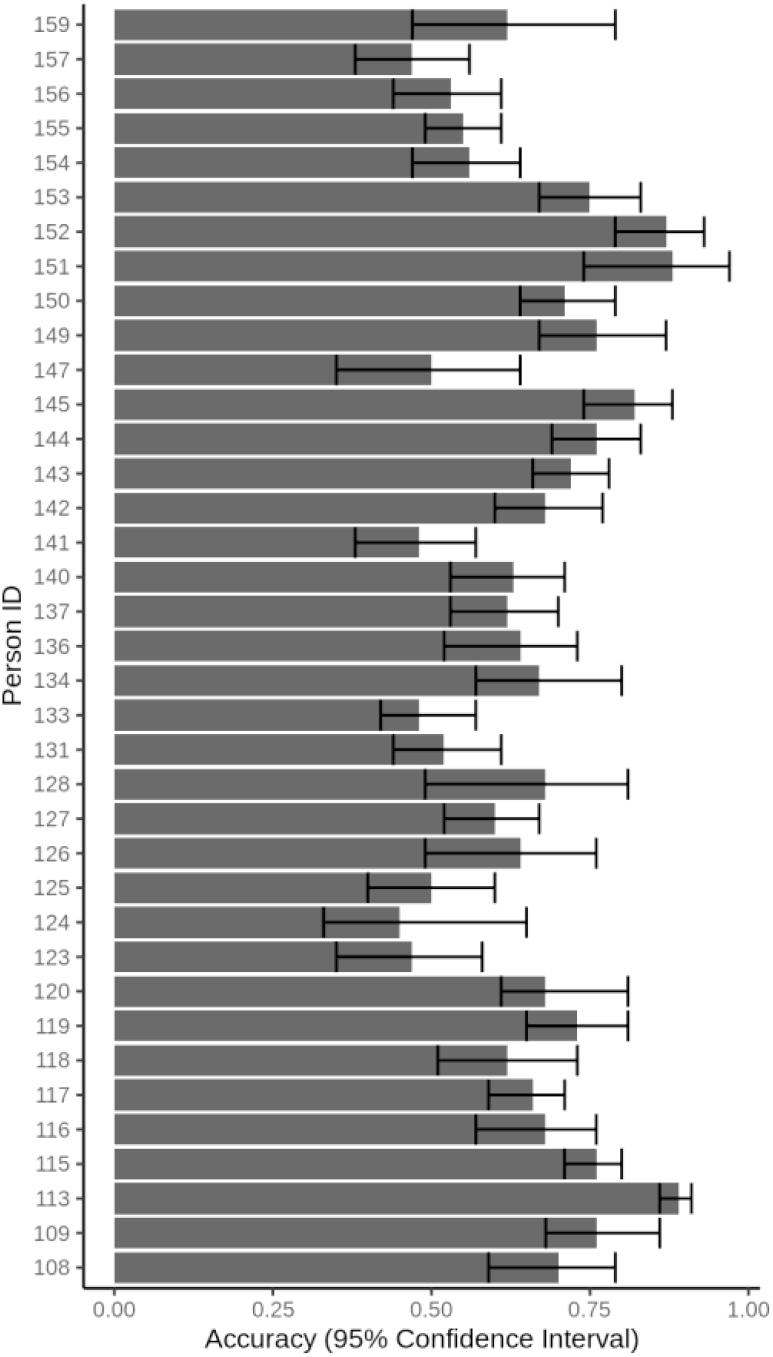
Participant-specific accuracy by Bayesian Additive Regression Tree: Posterior mean and 95% credible interval.

## Notes

### Competing Interest Statement

The authors have declared no competing interest.

