## Supplementary for "Quantifying the Uncertainty of Human Activity Recognition Using a Bayesian Machine Learning Method: A Prediction Study"

**Supplementary Digital Contents**

**Appendix 1:** Description of Bayesian Additive Regression Tree

Bayesian Additive Regression Trees (BART) is a nonparametric ensemble machine learning algorithm utilizing a sum of regression trees. Rather than relying on a single complex prediction model, BART combines multiple regression trees that provide good predictive performance as a whole. The trees are kept simple (allowed to have only a few branches) to prevent over-fitting, so that optimal out-of-sample prediction performance is achieved. To model our multinomial outcome containing six physical activity categories, we used the BART package in R software (1). Specifically, the package uses probit model to estimate the conditional probability of a response category *j* as:

$$P(y_{ij}|x_i) = \Phi\left(\mu_{ij} + f_j(x_i)\right),$$

for $j = \{1...K-1\}$, $K = 6$ outcome categories at *i*-th time point in accelerometer data. The function $\Phi$ represents the standard normal cumulative density function, with $\mu_{ij}$ representing intercept, and $f_j(x_i)$ representing the sum-of-tree function for category *j,* whose predictive features calculated from accelerometer signals at *i*-th time points are denoted $x_i$.

The general form of the ensemble BART function is represented as the sum of *m* trees as

$$f_j = g(x, T_1 M_1) + g(x, T_h, M_h) + g(x, T_m, M_m),$$

where $T_h$ represents tree sizes and $M_h$ represents the collection of leaf parameters as:

$$M_h = \left(\mu_{h1}, \mu_{h2}, \mu_{ht}, \ldots, \mu_{hB_h}\right),$$

for the total of $B_h$ leaves on the $h$ th regression tree.

As BART is a fully-Bayesian model, prior probabilities for these parameters need to be specified. Prior on tree depth imposes weak learning (trees with shallow depth), which is accomplished by assigning the probability of a node $d$ being non-terminal as:

$$\alpha(1+d)^{-\beta}, \alpha \in (0,1); \beta \in [0, \infty).$$

We used the recommended values of $\alpha = 0.95$ and $\beta$ = 2, such that the probability of a given tree having a complicated structure (large number of branches) is kept low (2).

The centered prior for the *t*-th leaf parameter covers a large probability range, ($\Phi[-3.0]$, $\Phi[3.0]$), and is specified as:

$$\mu_{ht} \sim N\left(0, \sigma_\mu^2\right)$$

$$\sigma_\mu = \frac{3.0}{k\sqrt{m}},$$

with larger *k* and *m* inducing a stronger shrinkage of the leaf parameters towards zero. We used *k=2* and *m=50* tees for our model as suggested (1).

To estimate the posterior distribution of parameter vector $y_{ij}$, we ran 2,000 burn-in iterations, followed by 2,000 samples for inference with thinning at every 100 iterations. From the three time-series of accelerometer signals that represent X, Y, and Z axis, we generated 58 predictive features described previously (3).

Accuracy metric of multi-category (as opposed to binary) prediction represents the proportion of agreement between the predicted and observed (i.e., ground truth) categories. We computed this category-specific accuracy metric for each of the 2,000 MCMC interactions, which were averaged over as the posterior mean of class-specific accuracy. Note that the predicted category for each time point was determined as the category with the highest estimated predicted probability, that is, $\hat{y}_i = \arg\max_j p\left(y_{ij}|x_i\right)$. For instance, the predicted category at *i*-th time point is $\hat{y}_i = 2$ when category-specific probabilities at this time point are (0.1, 0.5, 0.1, 0.1, 0.1, 0.1). It is possible to use these probability outputs from BART to calculate accuracy e.g., Brier score. However, we converted these probabilities into the categorical measure for comparison purpose, as the latter measure is the standard output from random forest.

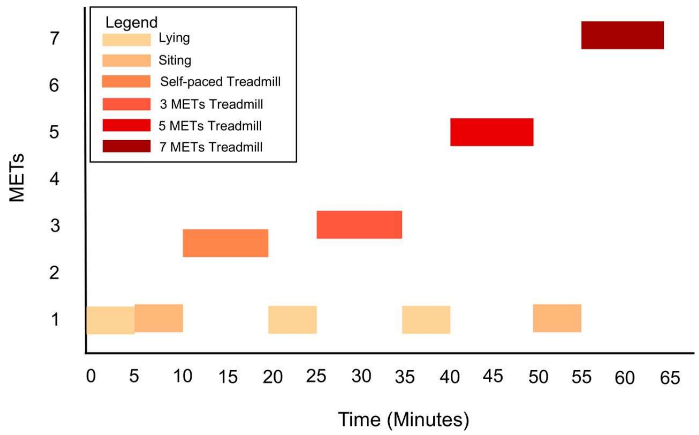


**Supplementary Figure 1.** Standardized activity protocol, illustrating the temporal transition of activity types and intensities performed by participants.
Abbreviation: METs, metabolic equivalents of task.

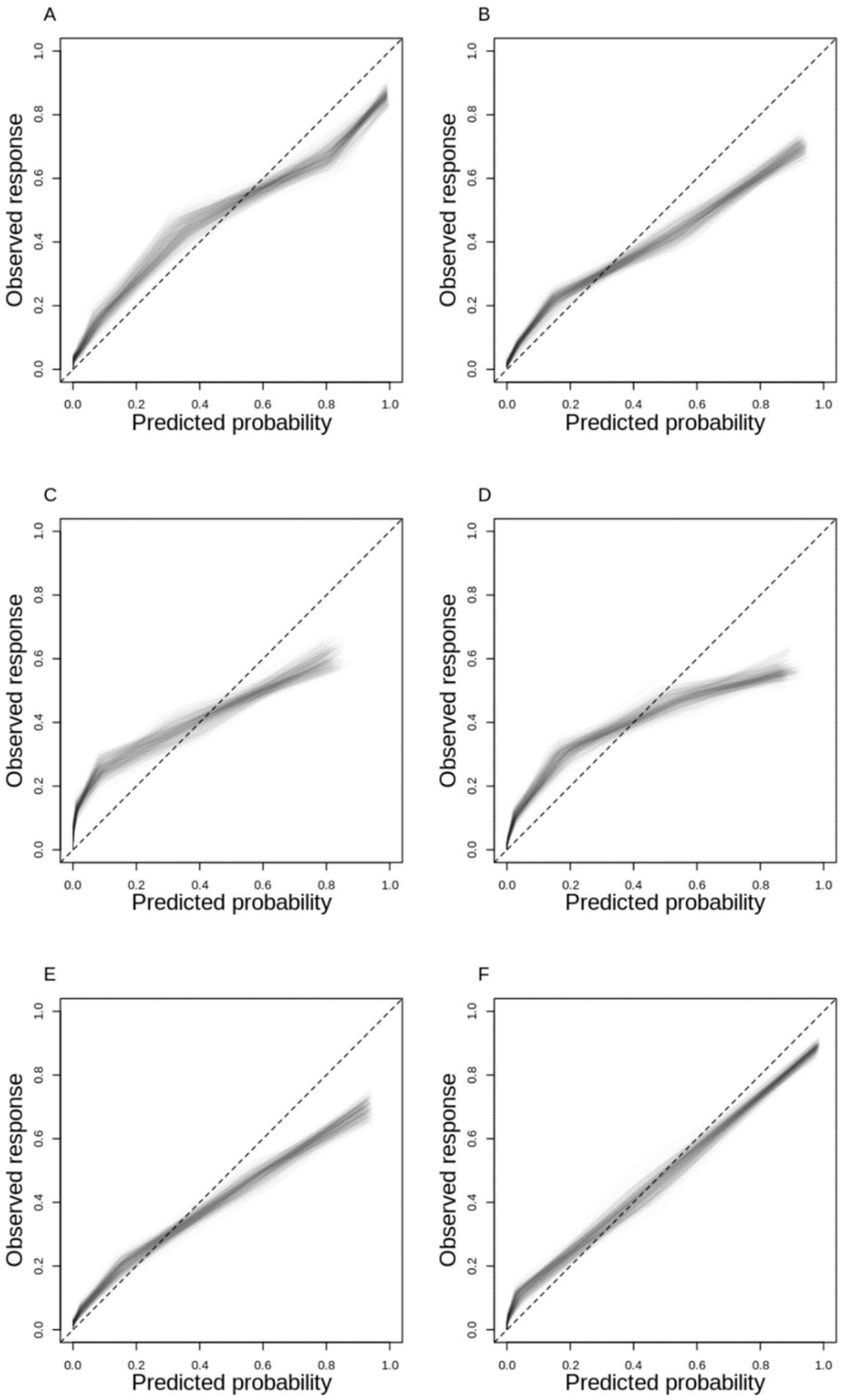


**Supplementary Figure 2 A-F**: Calibration plots of Bayesian Additive Regression Trees (BART) showing the goodness-of-fit between the decile of predicted probabilities and the proportion of outcomes that are: A) lying, B) sitting, C) self-paced walking, D) running at 3 METs, E) running at 5 METs, and F) running at 7 METs. Translucent lines represent the iterations of Markov Chain Monte Carlo.

The x-axis indicates the decile of predicted probability, and y axis indicates the proportion of the outcome corresponding to the deciles of predicted probabilities. Panel C and D shows a larger miscalibration (discordance) between the predicted and observed outcome probabilities, with BART underestimating the outcomes when predicting low probabilities, and overestimating when predicting high probabilities. On the other hand, BART generated predictive probabilities that closely match to the observed frequency when predicting Running 7 METs (Panel F).

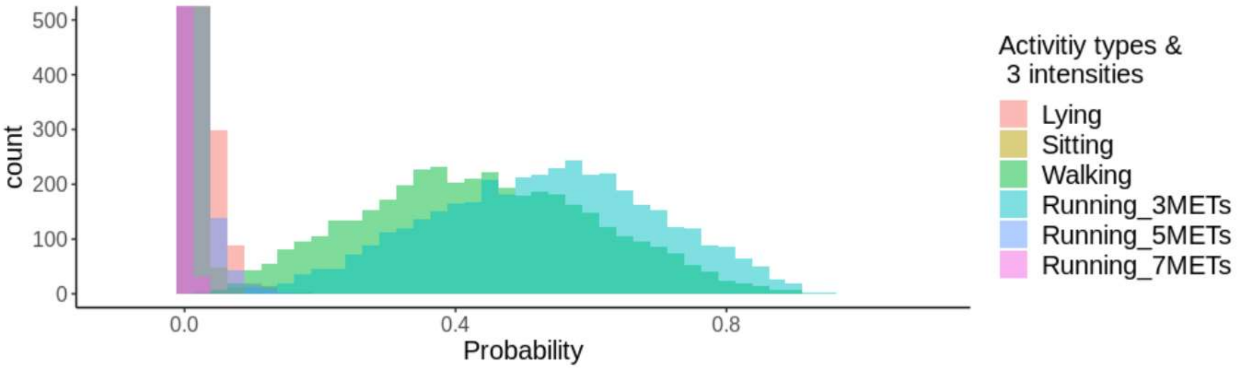


**Supplementary Figure 3.** Posterior predictive distribution of six activity categories at one time point, where the estimated distributions of walking and Running (3METs) are largely overlapping.

Abbreviation: METs, Metabolic Equivalents of Task.

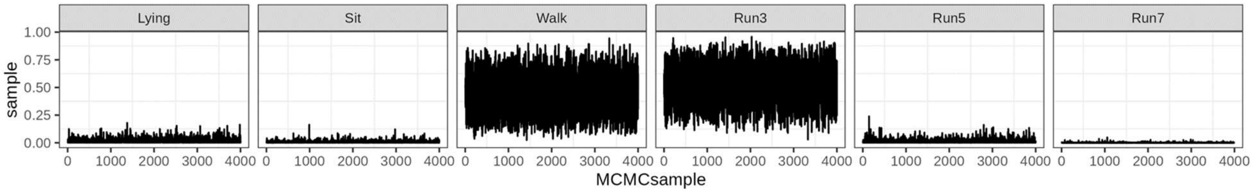


**Supplementary Figure 4.** Trace plots of a time point corresponding to Supplementary Figure 3 above, indicating the mixing of Markov Chain for each of the six activity categories.

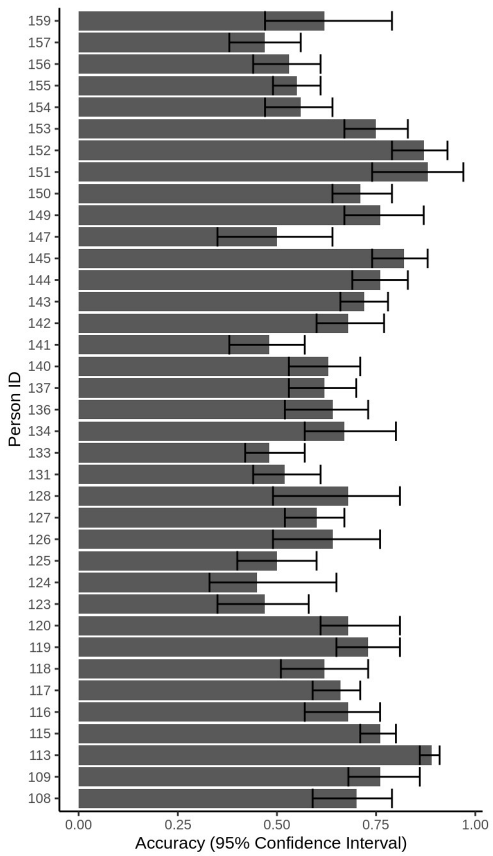


**Supplementary Figure 5.** Participant-specific accuracy by Bayesian Additive Regression Tree: Posterior mean and 95% credible interval.
